# Pregnenolone Reduces Provoked Craving and Cocaine Use in Men and Women with Cocaine Use Disorder: A Pilot Trial

**DOI:** 10.1101/2025.01.07.25320141

**Authors:** Elcin Sakmar, Stephanie Wemm, Nia Fogelman, Gretchen Hermes, Rajita Sinha, Verica Milivojevic

**Affiliations:** The Yale Stress Center, Department of Psychiatry, Yale University School of Medicine, New Haven, CT 06519

**Keywords:** pregnenolone, neuroactive steroids, cocaine use disorder, craving, cocaine use outcome

## Abstract

**Aim:** Chronic cocaine use is associated with decreases in neuroactive steroid levels. These adaptations may contribute to continued cocaine use and high relapse risk in individuals with cocaine use disorder (CUD). Thus, this pilot study assessed chronic treatment with 2 supraphysiologic doses of the neuroactive steroid precursor pregnenolone (PREG, 300 mg/day; 500 mg/day) to boost endogenous neuroactive steroid levels and assess its impact on provoked craving and cocaine use outcomes in an 8-week trial in men and women with CUD.

**Methods:** Fifty-five treatment-seeking individuals with CUD were randomly assigned to receive either placebo (PLA; n=18; 12M/6F), 300mg PREG/day (n=20; 15M/5F) or 500mg PREG/day (n=17; 12M/5F) for 8 weeks, along with outpatient weekly relapse prevention treatment. Plasma was collected at weeks 2, 5 and 7 to assess circulating pregnenolone levels. A subset of subjects participated in a 3-day experimental component of guided imagery exposure to stress, cocaine cue and neutral conditions in about week 2 of the trial to assess craving response. Cocaine use outcomes was also assessed over the 8-week treatment period. Intent-to-treat analyses were conducted using linear mixed effects models.

**Results:** There were no differences between treatment groups on demographic variables and baseline cocaine use. Plasma pregnenolone levels were higher in the 300mg and 500mg PREG groups compared to PLA (*p’s*< 0.032). Participant trial completion rates were 100% for PLA, 90% for 300mg and 94% for 500mg PREG groups. Placebo group had increased craving in response to stress (*p* < .001) and cocaine cue (*p* < .001) provocation, whereas the PREG groups showed no increased in provoked cocaine craving. For cocaine use outcomes during the 8-week trial, a significant main effect of treatment group (*p* = .005) on the weekly amounts of cocaine use showed e significantly lower amounts used in the 300mg PREG compared to the 500mg PREG group (*p* = 0.01) and to PLA (*p* = .047). There was also a trend for a treatment group main effect for days of cocaine used (p<.12)

**Conclusions:** These pilot findings suggest that supraphysiologic neuroactive steroid PREG doses reduces cocaine craving and may also improve cocaine use outcomes in treatment seeking individuals with CUD. Findings support further assessment and development of PREG in the treatment of CUD.

## Introduction

Cocaine use disorder (CUD) is a common and serious health problem (SAMHSA 2024) for which there are currently no approved medications. Early abstinence from cocaine is marked by dysregulated basal physiological and neuroendocrine tone, and stress- and cue-induced physiological, hypothalamic-pituitary-adrenal (HPA) axis and emotional changes, and significantly associated with increased drug craving, use and relapse risk (Sinha et al., 2006, Fox and Sinha, 2009, Back et al., 2005, Fox et al., 2008, Daughters et al., 2009, Sinha et al., 2003). This research has directly linked stress- and drug cue-induced craving, and their related disruption in stress biology to drug use and relapse risk suggesting that treatments that reverse these processes may be useful targets in drug relapse prevention (Milivojevic and Sinha, 2018).

Importantly, disrupted physiologic and neuroendocrine responses in substance use disorders (SUDs) are associated with reductions in GABAergic control of the stress axis (Biggio et al., 2007). Recent basic science and clinical evidence indicate that GABAergic neuroactive steroids (NAS) and their neuroactive precursor pregnenolone (PREG) normalize the HPA axis stress response and reduce drug craving, anxiety, and drug intake (Janak et al., 1998, Morrow et al., 2006, Besheer et al., 2010, Regier et al., 2014), suggesting that PREG may be acting by normalizing stress system upregulation and potentially decreasing related compulsive drug seeking. However, studies to develop NAS as potential medication targets in SUDs are rare. For example, the specific doses required for possible beneficial effects on craving, stress responsivity and drug use are unclear and the possible effects on other downstream NAS in humans are not fully understood.

Acute drug intake has been shown to increase GABAergic transmission and neuroactive steroid levels (Morrow et al., 2006, Morrow et al., 2020). Acute stress also increases levels of neuroactive steroids like ALLO (Torres et al., 2001), and pharmacological challenges of the HPA axis potently increase levels of PREG (Porcu et al., 2006). On the other hand, chronic drug use and related neuroadaptations down-regulate GABAergic transmission (Biggio et al., 2007) and decrease NAS levels in the brain and periphery (Purdy et al., 1991, Morrow et al., 2001, Morrow et al., 2006). This results in blunted drug-related NAS and GABA levels and altered stress responses (Biggio et al., 2007, Crowley and Girdler, 2014, Sarkar et al., 2011), that have been associated with greater emotion dysregulation and compulsive drug seeking (Porcu et al., 2016).

In the first set of human studies in addiction, we recently reported that in individuals with CUD and in those with alcohol use disorder (AUD) who received pregnenolone (PREG) daily doses of 300mg/day or 500mg/day or placebo, PREG doses decreased stress- and cue-induced craving, reductions in both stress- and cue-induced anxiety and normalizations in provoked autonomic responses (Milivojevic et al., 2022, Milivojevic et al., 2023). Importantly, preliminary evidence also suggests that levels of neuroactive steroids, such as pregnenolone, may be altered with chronic cocaine use (Milivojevic et al., 2019). These findings raise the question of whether potentiation of the neuroactive steroid system with its precursor pregnenolone may normalize chronic cocaine related adaptations in stress and reward pathways to decrease provoked craving and improve cocaine use outcomes.

Thus, the current pilot study assessed whether chronic treatment with two supraphysiologic doses of the neuroactive steroid precursor pregnenolone (PREG, 300 mg/day; 500 mg/day) would impact experimentally provoked craving in the laboratory and also show a positive signal for cocaine use and craving outcomes during an 8-week pilot trial in men and women with CUD. We hypothesized that pregnenolone at both doses would decrease craving and cocaine use over the treatment period compared to placebo.

## Methods

### Participants

Fifty-five treatment-seeking individuals (39 Male/16 Female) with cocaine use disorder (CUD) who responded to local advertisements around the New Haven area were enrolled between February 1, 2019, and April 30, 2023, and participated in the study (Fig. 1 CONSORT diagram). Inclusion criteria consisted of men and women 18-60 years old, with current CUD, ability to give written informed consent, and the ability to read and write English to complete study evaluations. Current CUD was determined using the Structured Clinical Interview for the Diagnostic and Statistical Manual of Mental Disorders 5 (SCID-5; (First et al., 2015). Exclusion criteria included: DSM-5 substance use disorder for any psychoactive substance, other than alcohol or nicotine, including opiate use disorder and including heroin (assessed and confirmed via urine toxicology screen in addition to SCID-5); any psychotic disorder or current Axis I psychiatric symptoms requiring specific attention; significant underlying medical conditions such as cerebral, renal, or cardiac pathology which in the opinion of study physician would preclude patient from fully cooperating or be of potential harm during the course of the study. Stable use of antidepressants and medications for medical conditions without known study drug interaction effects that did not preclude the participant from participating in the study as determined by the study physician were allowed to ensure generalizability of the study findings. All individuals underwent stringent medical assessments including electrocardiography and laboratory tests of renal, hepatic, pancreatic, hematopoietic and thyroid function, and a physical exam conducted by the study physician to determine study eligibility. Written and verbal consent was obtained from all participants and the study was approved by the Human Investigation Committee of the Yale University School of Medicine and was registered at ClinicalTrials.gov (NCT03953612).

**Figure 1.**
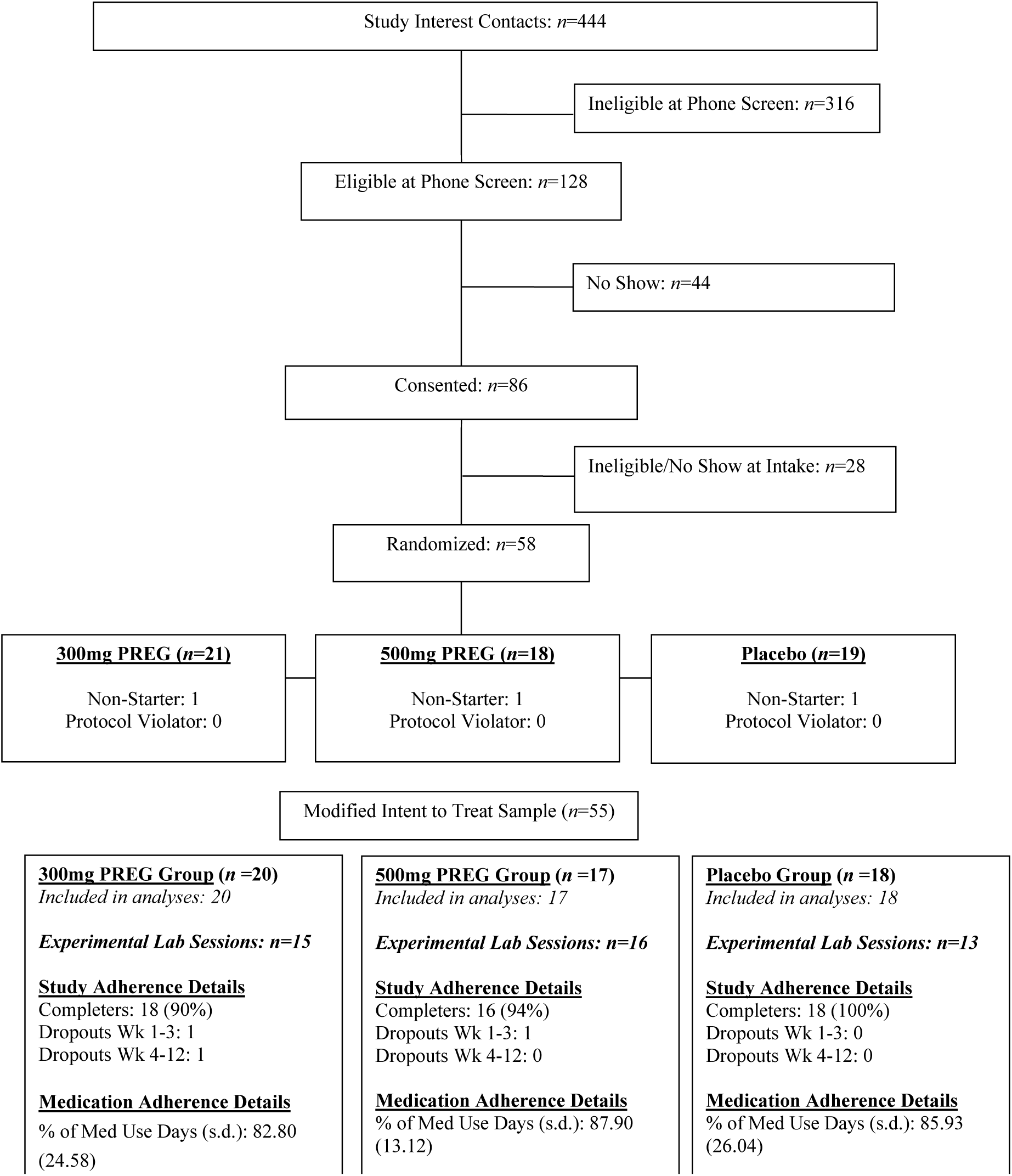
Study CONSORT Flow Diagram.

### Study Medication Randomization, Dosing and Adherence

Identical pregnenolone (150 mg and 250 mg strength for b.i.d. dosing) and placebo capsules that also contained 25 mg of Riboflavin were formulated by the Yale University research pharmacist (Investigational Drug Services, IDS) and prepared for dispensing in 1-week bottles. Participants took the study medication orally, twice per day in the morning and evening at 8AM and 8PM during the 8-week trial. Study participants and investigators were blind to the medication condition. Medication randomization was conducted by the Yale Stress Center Biostatistician and randomization was balanced for age, sex, smoking status, CUD severity, and education. Medication adherence during the trial was achieved in three ways: a) study medication pill counts, b) using the smartphone-based video monitoring tool eMocha Mobile Health, Inc. (Baltimore, MD), and c) blood levels of pregnenolone assessed repeatedly at approximately weeks 2, 5 and 7 of the trial (see Figure 1 and Figure 2).

**Figure 2.**
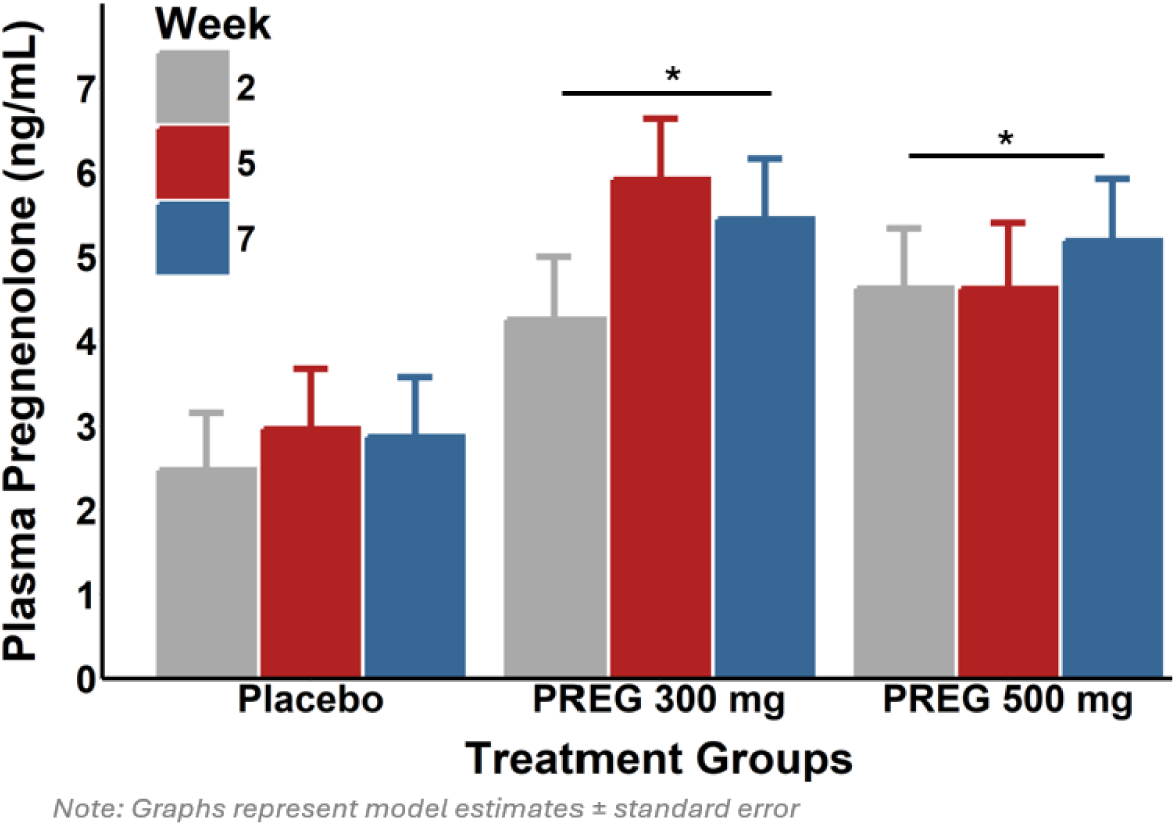
Pregnenolone levels as a function of treatment group and week in trial. A main effect of treatment group showed that the 300 mg PREG group and 500 mg PREG group both had higher pregnenolone levels than Placebo (*b* ± *SE:* 300 mg 5.2 ± 0.66, 500 mg 4.81 ± 0.66 v. Placebo 2.76 ± 0.63, *p*’s < 0.032).

### Procedures

After being deemed eligible for the study, participants were given the choice of taking part in either a fully outpatient treatment or an initial inpatient period followed by a transition to outpatient treatment for the remainder of the study period. Participants who opted for the inpatient-outpatient option were admitted to the Clinical Neuroscience Research Unit for a 2-week inpatient stay and were initiated on study medication on the inpatient unit. After completing the 2-week inpatient phase, subjects participated in the remaining 6-weeks of the treatment trial on an outpatient basis at the Yale Stress Center. A total of 9 participants took part in the inpatient option, see Table 1. Additionally, a subset of subjects (*n*=44; 80%of the sample) participated in an experimental component, where scripts for individualized guided imagery induction were developed (see below) using the well-established standardized procedures, as described in previous studies (Sinha, 2009, Sinha et al., 2009) during week 1 of their treatment. In week 2, a 3-day laboratory experiment involving participation in 3 separate experimental testing sessions on 3 separate days was conducted. Research staff were blind to order of imagery condition presented per day and subjects also remained blind until imagery presentation. Order of imagery condition was randomized and counterbalanced across subjects.

**Table 1.** Demographic Characteristics of the Sample.

|  | **Total**<br>**(N = 55)** | **Placebo**<br>**(n = 18)** | **300 mg**<br>**(n = 20)** | **500 mg**<br>**(n = 17)** | **Difference**<br>**Test** |
| --- | --- | --- | --- | --- | --- |
| Age | 47.27 (10.48) | 47.17 (9.44) | 48.05 (11.31) | 46.47 (11.06) | .751 |
| Years of Education | 12.47 (1.59) | 12.56 (1.58) | 12.50 (1.40) | 12.35 (1.87) | .551 |
| Years of Cocaine Use | 18.68 (12.08) | 14.36 (9.83) | 24.46 (12.06) | 16.80 (12.79) | .107 |
| Gender (Male) | 39 (70.91%) | 12 (66.67%) | 15 (75%) | 12 (70.59%) | .931 |
| Race |  |  |  |  | .374 |
| Caucasian | 19 (34.54%) | 8 (44.44%) | 5 (25%) | 6 (35.29%) |  |
| African American | 35 (63.64%) | 9 (50%) | 15 (75%) | 11 (64.71%) |  |
| Other | 1 (1.82%) | 1 (5.56%) | 0 | 0 |  |
| # of Smokers (YES) | 44 (80%) | 16 (88.89%) | 14 (70%) | 14 (82.35%) | .370 |
| Lifetime Mood Disorder | 11 (20%) | 2 (11.11%) | 5 (25%) | 4 (23.53%) | .488 |
| Lifetime Anxiety Disorder (incl. PTSD) | 17 (30.91%) | 5 (27.78%) | 8 (40 %) | 4 (23.53%) | .504 |
| Current Meds# (during the study period) |  |  |  |  |  |
| Mood & Anx. Dis. | 7 (12.73%) | 3 (16.67%) | 2 (10%) | 2 (11.76%) | .883 |
| Other | 40 (72.73%) | 14 (77.78%) | 14 (70%) | 12 (70.59%) | .864 |
| Inpatient (YES) | 9 (16.36%) | 3 (16.67%) | 3 (15%) | 3 (17.65%) | 1 |

*Note.* Each cell shows mean (sd) or frequency (%).

### Imagery Script Development Procedures

Imagery script development was conducted in week 1 in a session prior to the laboratory experiment. Procedures are based on methods developed by Lang and his colleagues (Lang et al., 1983), and further adapted and validated in our previous studies (Sinha et al., 2009, Sinha et al., 2003, Sinha et al., 2000, Sinha et al., 1999, Fox et al., 2005). Briefly, the stress imagery script was based on subjects’ descriptions of a recent “most stressful” adverse personal event that made them “sad, mad or upset”, that they were not able to control in the moment. “Most stressful” was determined by having the subjects rate their perceived stress on a 10-point Likert scale where 0 = not at all stressful and 10 = the most stress they felt recently in their life. Only situations rated as 8 or above were accepted as appropriate for script development (e.g., being fired from their job, marital conflict situation). The cocaine-related cue scripts were developed by having subjects identify a recent situation that included cocaine-related stimuli and resulted in subsequent cocaine use. Cocaine related situations that were associated with negative affect or psychological distress were not allowed. A relaxing, non-physiologically arousing and non-cocaine related script was developed from the subjects’ description of a personal, relaxing situation (e.g., being at the beach; fall afternoon reading at the park). In addition to the script development, on the day of the first laboratory session, subjects were brought into the testing room in order to acclimatize them to specific aspects of the study procedures including the subjective rating forms and training in relaxation and imagery procedures, as previously described in (Sinha et al., 2009).

### Laboratory Sessions (Conducted across Three Separate Days)

Participants were instructed to abstain from using cocaine after midnight prior to the laboratory sessions. Female subjects completed the laboratory sessions during the follicular phase of their menstrual cycle when both progesterone and estradiol levels are low and remain stable. Participants were brought into the testing room at 1:30 p.m., after a standard lunch provided at 1:00 p.m. after self-administration of study drug. Patients who were smokers were allowed a smoke break immediately prior to 1:30 p.m. in order to reduce potential nicotine withdrawal during the session. After settling in a sitting position on a reclining chair in an experimental testing room, a heparin-treated intravenous (IV) catheter was inserted at 2:00 p.m. by the research nurse in the antecubital region of the subject’s non-preferred arm in order to periodically obtain blood samples. A Critikon Dinamap 120 Patient Monitor was also placed on the subject’s preferred arm, including a pulse sensor which was placed on the subject’s forefinger. This was followed by a 40-min adaptation period during which the subjects were provided relaxation instructions to ensure stable psychophysiological state prior to each lab day. Immediately following the adaptation period, subjects were provided headphones and given the following instructions for the 5-min imagery procedure: “Close your eyes and imagine the situation being described, ‘as if’ it were happening right now. Let your body and mind get completely involved in the situation, doing what you would do in the real situation”. Cocaine craving was assessed at various timepoints, including prior to imagery (baseline timepoint 0, +45, +65), immediately following imagery presentation (+77), and every 15 min after the imagery period, up to 75 min (+90, +105, +120, +135, +150.

#### Behavioral Counseling

Weekly behavioral counseling was provided to all participants during the 8-week treatment period, using the empirically validated, standardized 12-Step Facilitation Therapy Manual, which included encouragement to attend self-help meetings and addressed triggers for relapse prevention (Nowinski, 1992).

#### Safety measures

At each weekly visit, side effects were assessed using the Systematic Assessment for Treatment Emergent Events (SAFTEE) (Levine and Schooler, 1986).

### Primary and Secondary Outcomes

#### Primary Laboratory Outcome: Provoked Cocaine Craving

The desire for using cocaine in the laboratory sessions was assessed using a 10-point visual analog scale (VAS) in which 0 = “not at all” and 10 = “extremely high”.

#### Secondary Outcomes

##### Pregnenolone Levels

Blood samples were collected approximately in weeks 2, 5 and 7 to measure plasma pregnenolone levels. All plasma blood collection tubes were centrifuged in a cold centrifuge (4C) and placed on ice immediately upon centrifugation. Plasma was then aliquoted into cryovial tubes and stored at −80C until processing at the Yale Center for Clinical Investigation Molecular Core Laboratories using a commercially available pregnenolone Enzyme-Linked Immunosorbent Assay (ELISA) kit (Eagle Biosciences, Inc., Nashua, NH). The ELISA kit has a sensitivity of 0.05 ng/mL, and a 100% specificity for pregnenolone. The coefficients of variation (CV) of intra-assay and inter-assay were <10.6% and <14.5%, respectively. For this ELISA, the cross reactivity between pregnenolone and other steroids was 6% for progesterone, 4.7% for 5alpha-androstanediol, 0.4% for pregnenolone sulfate, 0.3% for androstanedione, 0.2% for DHEAS, and less than 0.1% for several other steroids (e.g. androsterone, aldosterone, androstenedione, cholesterol, corticosterone, 5alpha-DHT, 17beta-estradiol, testosterone).

##### Cocaine Use Data

Participants completed daily diary data prompts on their phone, indicating the quantity of cocaine consumption that day and the day prior. Additionally, participants completed the Substance Use Calendar using the Timeline FollowBack method providing daily cocaine and other drug use retrospectively each week during the treatment period (Sobell and Sobell, 1992) when attending check in visits at the Yale Stress Center per study protocol. Daily cocaine use data was finalized using the most proximal data (daily prompts on use for that day). If this was unavailable due to non-completion of the daily prompt, then prior cocaine use days from the next prompt was used. If both were missing, daily Substance Use Calendar data collected on a weekly basis was filled in.

#### Other Related Outcome

##### Baseline and Study Period Cocaine Craving

Cocaine craving at baseline (pre study) and weekly during the study period was assessed using the Cocaine Craving Questionnaire (CCQ)-NOW, a brief well validated 10-item self-report craving scale (Tiffany et al., 1993).

### Data and Statistical Analysis

#### Power analysis

Sample size and power analysis determinations were based on our initial findings on the effects of progesterone-stimulated ALLO on cocaine craving (Milivojevic et al., 2016) to generate effect sizes and sample sizes. The effect size “f” was estimated using the formula (Cohen, 1988) f= sq root of (dfnum X F/N). We obtained an effect size of f=.39 for provoked cocaine craving and at two-tailed α=.05 and power (1-β)=0.80 and using Cohen (1988) page 384 a sample size of 14 per group was found to be adequate for testing of medication effects on experimentally provoked cocaine craving. An additional goal of the study was to generate preliminary data on cocaine use outcomes data to determine effect sizes for a larger study. Based on the preliminary power analysis detailed above, recruitment goal for the current study was 20 participants per group, which would ensure at least 14 individuals in each group after attrition and was deemed adequate for the current pilot study.

#### Data Analysis

R Studio 2023.09.1 +494 was used for data cleaning, analysis, and visualization (Packages: car v.3.1-3, emmeans v.1.10.5, DHARMa v.0.4.7, ggplot2 v.3.5.1, ggpubr v.0.6.0, MASS v.7.3-60, psych v.2.4.6.26). Kruskal Wallis test for continuous variables and Fisher’s Exact test for categorical variables were used to investigate treatment groups differences on the demographic characteristics (Table 1), adverse events (Table 2), and medication compliance during the study (Figure 1). Negative binomial regression was used to compare treatment groups in terms of amount of cocaine use ($), percentage of days participants used cocaine, and cocaine craving (Table 3) for both baseline and the study period. To calculate the average amount of cocaine use per period (i.e., baseline and study period, separately), the sum of daily amount of cocaine use was divided by the total number of days recorded. Then, the percentage of days participants used cocaine was the total days when cocaine was consumed during the assessment period divided by the total number of days recorded and multiplied by 100. Additionally, to calculate the average cocaine craving during the study period, the sum of weekly cocaine craving scores was divided by the total number of weeks recorded. Moreover, cocaine craving during the imagery lab sessions was assessed at timepoints 0, 45, 65 (baseline), 77 (imagery) and 90, 105, 120, 135, and 150 (recovery). Linear mixed effects models were conducted to investigate whether there was a Group by Time by Lab Condition (Neutral, Stress, Cue) interaction with a random intercept per participant. Control variables that were modeled in all analyses were gender (0=female), inpatient (0=outpatient), and age. Pregnenolone levels (plasma) during the trial were also assessed at about Weeks 2, 5, and 7 of the 8-week trial period. Linear mixed effects models were employed to examine whether there was a group (Placebo, 300 mg, 500 mg) by Time (Week 2, 5, and 7) interaction with a random intercept per participant.

**Table 2.** Frequency of adverse events occurring in ≥5% of participants in either treatment group.

| **Adverse Event** | **Placebo**<br>**(n = 18)** | **300mg PREG**<br>**(n = 20)** | **500mg PREG**<br>**(n = 17)** | ***p-values*** |
| --- | --- | --- | --- | --- |
| Pain/Injury | 2 | 5 | 3 | .563 |
| Headache | 3 | 2 | 2 | .883 |
| Infection/Immune System | 1 | 2 | 3 | .574 |
| GI | 1 | 1 | 1 | 1 |
| Inflammation/Allergy | 1 | 1 | 1 | 1 |
| Sleep Disturbance | 1 | 0 | 2 | .194 |
| Fatigue | 1 | 0 | 1 | .529 |
| Suicidal Thoughts | 1 | 0 | 0 | .636 |
| Lightheadedness | 0 | 1 | 0 | 1 |

Note: Multiple occurrences of the same adverse event for a participant were counted once in the frequency of the adverse event.

**Table 3.** Baseline and Study Period Cocaine Use & Craving.

|  | **Total** | **Placebo** | **300 mg** | **500 mg** | ***p*-value** |
| --- | --- | --- | --- | --- | --- |
| **<u>Baseline</u>** |  |  |  |  |  |
| Cocaine Amount (\$) | 38.21 (35.79) | 34.89 (38.32) | 35.28 (32.26) | 45.25 (37.89) | .432 |
| % of Use Days | 53.35 (31.01) | 47.61 (26.88) | 48.28 (31.43) | 65.50 (33.20) | .266 |
| Cocaine Craving | 40.09 (17.48) | 42.27 (16.71) | 35.75 (17.44) | 42.71 (18.61) | .576 |
| **<u>Study Period</u>** |  |  |  |  |  |
| Cocaine Amount (\$) | 21.81 (30.10) | 28.28 (43.99) | 12.3 (17.57) | 26.44 (20.59) | .005 |
| % of Use Days | 27.31 (22.88) | 28.61 (26.19) | 19.9 (20.46) | 35.12 (20.05) | .122 |
| Cocaine Craving | 27.92 (11.35) | 27.83 (13.04) | 28.50 (11.58) | 27.37 (9.65) | .963 |

*Note.* Each cell shows mean (sd).

## Results

### Participants’ Baseline Characteristics

Baseline characteristics of the modified intent-to-treat sample are summarized in Table 1. The three medication groups did not differ significantly in any of the demographic variables or in baseline amount and frequency of cocaine use or baseline cocaine craving using the CCQ-Brief, see Table 3.

#### List of Mood & Anx. Dis. Meds Participants Reported

Bupropion, Escitalopram, Fluoxetine, Mirtazapine, Trazodone

#### List of Other Meds Participants Reported

Acetaminophen, Albuterol, Allergy Medication, Amlodipine, Amoxicillin, Aspirin, Atorvastatin, Azithromycin, Benzonatate, Bismuth Subsalicylate, Brimonidine, Cannabis, Celecoxib, Cetirizine, Chlorothiazide, Chlorthalidone, Ciprofloxacin, Codeine, Cyclobenzaprine, Diclofenac Sodium, Diphenhydramine, Doxylamine, Ferrous Sulfate, Gabapentin, Generic Cough Medicine, Hydrochlorothiazide, Ibuprofen, Iron Supplement, Lamotrigine, Levothyroxine, Lidocaine, Lisinopril, Loratadine, Lotensin, Maalox, Methimazole, Metoprolol, Metronidazole, Nabumetone, Naproxen, Oxycodone, Pantoprazole, Pepsin, Polyethylene, Glycol, Prednisone, Propranolol, Ramelteon, Ranitidine, Ropinirole, Rosuvastatin, Sinus Medication, Theraflu, Topiramate, Tramadol, Trimtone

### Study and Medication Adherence

Study completion rates included 18/18 (100%) completers in the PLA group, 18/20 (90%) completers in the 300mg PREG group and 16/17 (94%) completers in the 500mg PREG group. The three groups did not differ in medication adherence, including number of medication doses participants used (*χ²*(2) = 0.07, *p* = .966), and percentage of days on which any study med was taken (*χ²*(2) = 0.7, *p* = .704), see Figure 1, Consort Diagram.

### Safety and Adverse Events

Serious adverse events (SAE’s) only occurred in one participant in the 300mg PREG group, who was briefly hospitalized for ongoing lightheadedness that was pre-existing. The proportion of participants who reported at least one non-serious adverse events (AEs) during the trial was not significantly different between the PREG groups and the placebo group (see Table 2 for side effects profile).

### Pregnenolone Levels

Plasma pregnenolone levels by treatment group and week indicated a main effect of Week (*F*(2, 71) = 3.32, *p* < 0.042) and Treatment Group (*F*(2, 44) = 4.14, *p* < 0.023). Overall, Week 5 and Week 7 levels were higher than Week 2 (*p*’s < 0.034) and levels in the 300 mg and 500 mg PREG groups were higher than Placebo (*p*’s < 0.032), see Figure 2.

### Cocaine Craving (Provoked in the Lab)

The results showed a significant interaction between treatment group and lab condition (*F*(4) = 3.50, *p* = .007). Participants in placebo group reported higher cocaine craving in response to stress (*p* < .001) and cue (*p* < .001) conditions compared to neutral condition, while the 300 mg and 500 mg PREG groups did not show such increases in stress and cue-provoked craving compared to the neutral condition (*p* = n.s.) (Figure 3).

**Figure 3.**
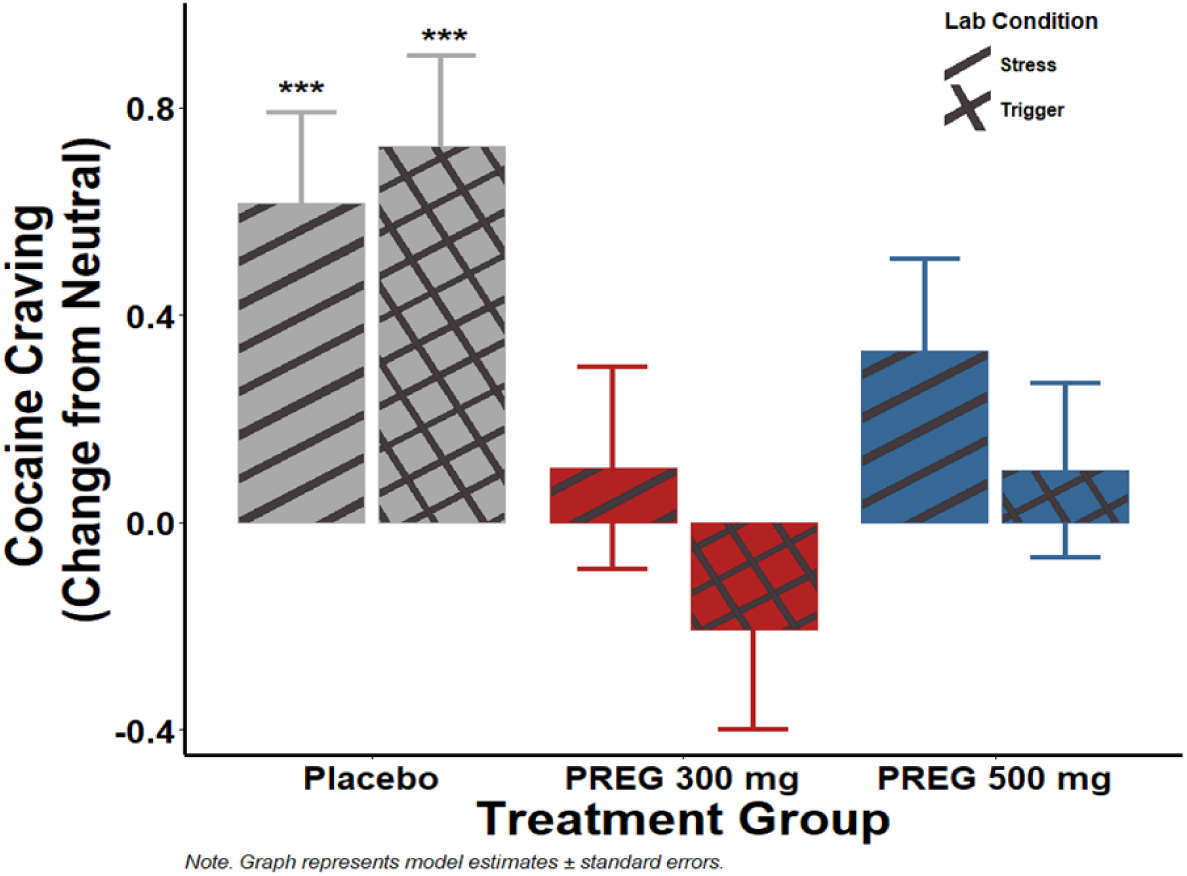
Provoked laboratory cocaine craving as a function of imagery condition and treatment group. Cocaine craving was higher in response to stress (*p* < .001) and cue (*p* < .001) conditions compared to neutral condition in the placebo group, but not in the PREG groups (*p* = n.s.).

### Treatment Period Cocaine Use

Treatment groups significantly differed on the average amount of cocaine use ($) during the study period (*χ²*(2) = 10.49, *p* = .005). The 300 mg group reported significantly lower amounts of cocaine use compared to the Placebo (*p* = .047) and 500 mg (*p* = .001) groups (Table 3, Figure 4). There was also a trending treatment group main effect for the percentage of days participants used cocaine (*χ²*(2) = 4.20, *p* = .122).

**Figure 4.**
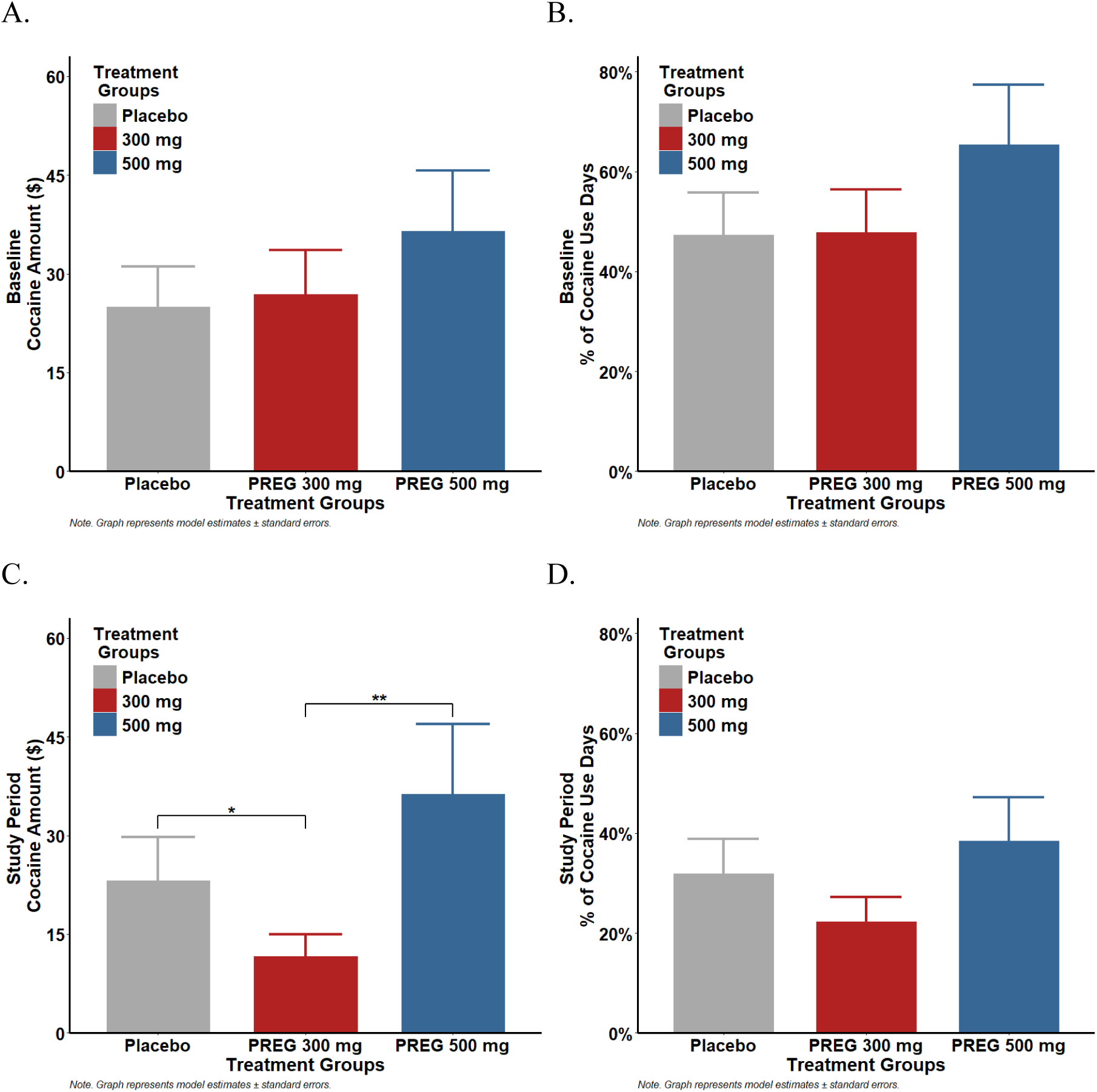
Baseline and study period cocaine use outcomes. Baseline (pre-study) average amount of cocaine use ($) (A) and percentage of days participants used cocaine (B) did not differ between the treatment groups. (C) Treatment groups significantly differed on the average amount of cocaine use ($) during the study period (*χ²*(2) = 10.49, *p* = .005), where the 300 mg PREG group reported significantly lower amounts of cocaine use compared to the Placebo (*p* = .047) and 500 mg PREG (*p* = .001) groups. (D) There was a trending treatment group main effect for the percentage of days participants used cocaine (*χ²*(2) = 4.20, *p* = .122

### CCQ-Brief Cocaine Craving During Study Period

Treatment groups did not significantly differ on weekly CCQ-Brief cocaine craving during the study period (*χ²*(2) = 0.08, *p* = .963), see Table 3.

## Discussion

To our knowledge, this is the first clinical trial that investigated two doses of pregnenolone to reduce cocaine use outcomes in treatment seeking men and women with cocaine use disorder (CUD). In this pilot study, twice-daily doses of 300mg and 500mg pregnenolone were found to be safe, well tolerated and with excellent medication adherence and study completion rates. Positive findings on primary outcome were observed with treatment-related reductions in stress- and cue-induced cocaine craving and significant maintained increases in plasma pregnenolone levels in PREG dose groups compared to placebo, Furthermore, secondary cocaine use outcomes showed dose-specific reductions in amount of cocaine use and trending level for days of cocaine used for the PREG 300 mg dose during the eight-week treatment period compared to PREG 500 mg/day and to placebo. These findings suggest pregnenolone 300 mg/day may reduce provoked cocaine craving and cocaine use outcomes and needs further investigation in a larger study.

Twice daily, oral pregnenolone significantly increased circulating levels of pregnenolone at both the 300mg and 500mg pregnenolone doses compared to placebo. The levels were assessed repeatedly throughout the trial at approximately weeks 2, 5 and 7 and showed significantly increased blood levels that remained stable throughout the study period. While the current trial implemented a twice daily dosing in the morning and evening in order to achieve stable increased blood levels, it was still not known whether chronic administration of pregnenolone in humans would potentially have poor bioavailability, rapid metabolism, and instability in the body (Vallee, 2016), which could limit its clinical efficacy. In order to address these concerns, we assessed repeated plasma pregnenolone levels over three days in a subset of the participants from the current trial, who were admitted on an inpatient unit for the first two weeks of pregnenolone administration (Gao et al., 2024). Repeated blood sampling in week two of chronic pregnenolone administration showed that both 300 mg/day and 500 mg/day achieved stable and reliable elevated plasma pregnenolone levels over 32.5 hour period in individuals with CUD, supporting good bioavailability of pregnenolone in these samples.

A large subset of participants in the current study also volunteers for the experimental provocation laboratory procedure conducted in week 2 of treatment, in which repeated assessments of stress- and cue-provoked cocaine craving was made. Both doses of pregnenolone treatment was found to decrease stress- and cocaine cue-induced craving compared to the placebo group, consistent with our prior preliminary report (Milivojevic et al., 2022) and with effects seen in those with alcohol use disorder (Milivojevic et al., 2023).

Given the 8-week trial, we were able to preliminarily assess dose-specific reductions in cocaine use in the current study. All three groups displayed reductions in the cocaine amount used as well as the percentage of days on which cocaine was used compared to the baseline period. However, the 300mg PREG group additionally had significantly lower amounts of cocaine used during the treatment period compared to both the 500mg PREG and to placebo groups. While the current study cannot elucidate pregnenolone’s specific mechanism of action underlying the observed benefits, previous pre-clinical studies which showed that pregnenolone reduces alcohol intake (Besheer et al.,2010), and clinical experimental studies showing that pregnenolone reduces stress and cue-induced craving and anxiety, and normalizes critical components of the sympathetic nervous system (Milivojevic et al., 2022, 2023), support the findings of reductions in cocaine use observed in the current study.

In the current study, we also assessed weekly self-report of cocaine craving, and while we didn’t see any main effects of pregnenolone this could be due to the fact that craving decreased significantly in all the groups during the treatment period. This reduction in overall craving could also be explained by the weekly individual counseling that all participants received during the entire treatment period.

Overall, this pilot clinical study is the first to test the effects of supraphysiologic doses of oral pregnenolone and show that the treatment was well-tolerated and safe. This is consistent with previous clinical trials using pregnenolone up to 500mg per day in patients with schizophrenia and bipolar disorder (Marx et al., 2009, Marx et al., 2014, Brown et al., 2014). We also didn’t observe any sex differences in any of the outcomes, suggesting that pregnenolone is safe and effective in both men and women with cocaine use disorder.

This pilot clinical trial had considerable strengths such as excellent retention and completion rates, sustained elevated levels of plasma pregnenolone throughout the trial, and recruitment of both men and women with approximately one third of the sample being female. However, some important limitations include that this is indeed a pilot study with initial findings, so it is critical that this should be replicated in a larger clinical trial with a longer treatment period and a follow up period to further assess the effects of pregnenolone on cocaine use outcomes.

## Data Availability

All data produced in the present study are available upon reasonable request to the authors.

## Acknowledgements

This study was supported by the National Institutes of Health (NIH), grants K01-DA046561 (Milivojevic) from the National Institute on Drug Abuse (NIDA) and R01-AA030923 (Milivojevic) and R01-AA026514 (Sinha) from the National Institute on Alcohol Abuse and Alcoholism (NIAAA). The authors declare no conflict of interest.

